# Elementary time-delay dynamics of COVID-19 disease

**DOI:** 10.1101/2020.03.27.20045328

**Authors:** José Menéndez

## Abstract

An elementary model of COVID-19 dynamics—based on time-delay differential equations with a step-like survival function—is shown to be in good agreement with data from China and South Korea. The time-delal approach overcomes the major limitation of standard Susceptible-Exposed-Infected-Recovered (SEIR) models based on ordinary differential equations, namely their inability to predict the observed curve of infected individuals as a function of time. The model is also applied to countries where the epidemic is in earlier stages, such as Italy and Spain, to obtain estimates of the total number of cases and peak number of infected people that might be observed.

---

The urgency to develop reliable mathematical models of the COVID-19 outbreak has become apparent over the past few weeks. While advanced network dynamics approaches have the potential to address the spatio-temporal dynamics in the most realistic way [1], simpler deterministic compartmental models have the advantage that they can be fitted to present data to predict the near-term impact in any given country. A conceptual ‘Susceptible-Exposed-Infectious-Removed’ (SEIR) framework has been proposed by Lin *et al*. and applied to earlier Wuhan data [2]. Several other SEIR variants are being proposed on a daily basis [3,4]. However, significant limitations of these models are beginning to emerge as more data from China and South Korea becomes available. Tabulations of data for each country contain the cumulative number of cases *C* ^*^ (*t*) and current number of infected people *I* ^*^ (*t*) [5]. The difference *R*^*^ (*t*) = *C* ^*^(*t*) − *I* ^*^ (*t*) gives the cumulative number of recovered and deceased patients. Both China and South Korea are past the time *t*_*peak*_ at which *I* ^*^(*t*) reaches its maximum. An important characteristic of the data is that the ratio *R*^*^ (*t*_peak_) / *C* ^*^ (*t*_peak_) is very small for both countries (20% in the case of China and 5% in the case of South Korea). To reproduce these observed ratios within SEIR models, very long mean infection periods are required, exceeding 100 days. Such long infection periods are unrealistic and lead to a t > *t*_*peak*_ decrease in the number of infected people that is much slower than observed.

The discrepancy between SEIR models and the available data becomes even worse if one considers the fact that the time lag for reporting a recovered person is likely to be much shorter than the incubation time for the disease. Therefore, the “instantaneous” predicted quantities *C*(*t*) and *R*(*t*) from the models do not correspond to the observed *C* ^*^ (*t*) and *R*^*^ (*t*) but to “corrected” quantities that can approximated as

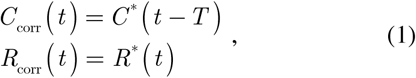

where *T* is the mean incubation time. Using *T* = 7 days, in line with a reported incubation time between 5 and 11 days [6], one obtains the data in Fig. 1 and Fig. 2. In the case of China, for example, the 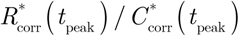 has ratio now become as small as 8%.

**FIG. 1.**
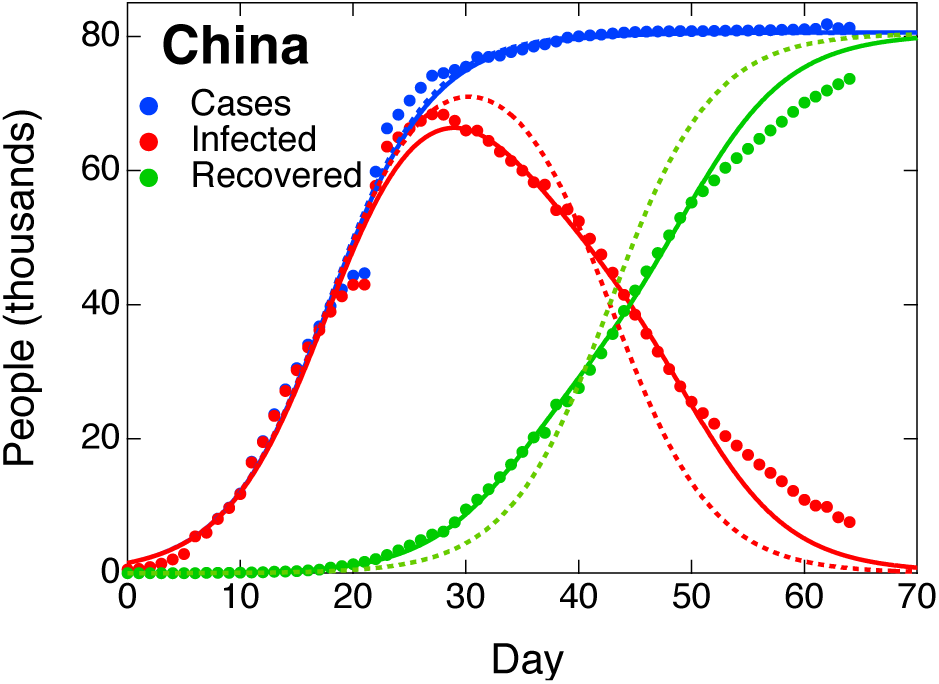
COVID-19 in China: blue dots show the number of cumulative cases, red dots the number of infected people at any given time, and green dots the cumulative number of recovered individuals. The data have been corrected according to Eq. (1) using *T* = 7 days. Day 0 is January 15, 2020. The dotted lines of the same color represent the prediction from Eq. (2) with parameters from Table I. The solid line is the prediction using Eq. (5).

**FIG. 2.**
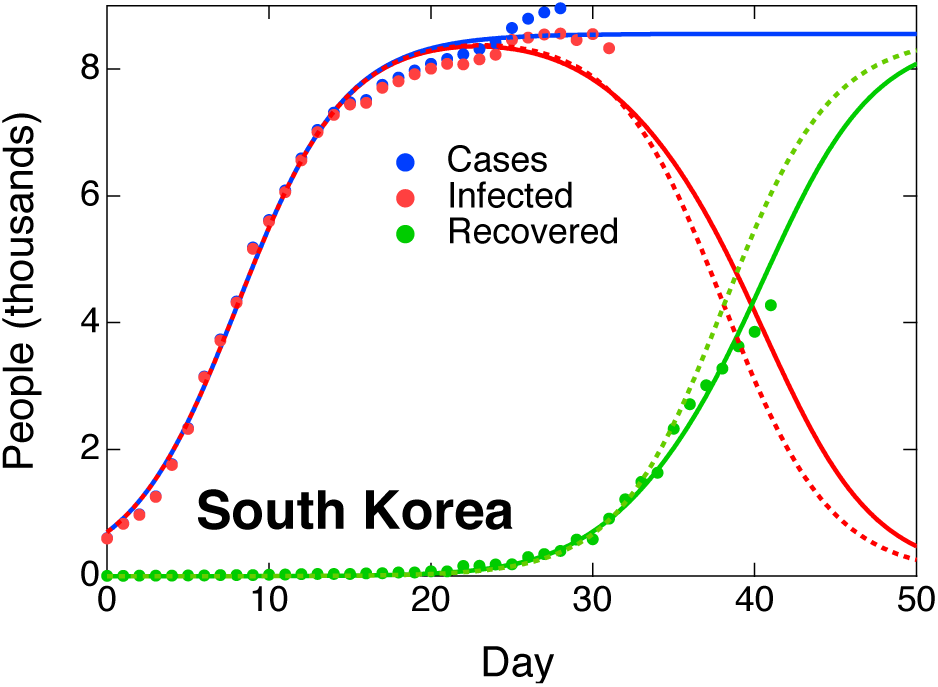
COVID-19 in South Korea: blue dots show the number of cumulative cases, red dots the number of infected people at any given time, and green dots the cumulative number of recovered individuals. The data have been corrected according to Eq. (1) using *T* = 7 days. Day 0 is February 16, 2020. The dotted lines of the same color represent the prediction from Eq. (2) with parameters from Table I. The solid line is the prediction using Eq. (5).

The qualitative discrepancy between SEIR models and actual data suggests that the SEIR ordinary differential equations must be replaced by delay differential equations that explicitly take into account the fact that people who recover at time *t* became sick at earlier times. In this paper, a very simple time-delay model is proposed that gives a reasonably good account of the China and South Korea and data.

The first approximation in the simplified time-delay model presented here is to ignore the “Exposed” category in SEIR models and start from a simpler SIR model [7]. An SEIR extension would be straightforward but may not be warranted at this time in view of the uncertain quality and consistency of the available data. Furthermore, it is assumed that the total number of people affected by the disease remains at all times much lower than the total country population, which makes it possible to eliminate the “susceptible” category *S* as well. This leads to an “IR” model which can be written as

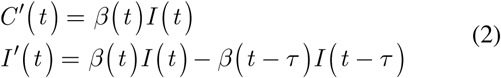

Here *β*(*t*) is the transmission rate—assumed to be time dependent—, and *τ* the mean convalescence time. The equations imply a step-like survival function in time-delay epidemiological models [8].

A necessary refinement of any model that attempts to reproduce the COVID-19 dynamics in China and South Korea is the incorporation of societal protective measures. Lin *et al*. [2] proposed

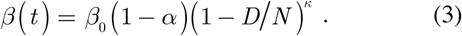

where *β*_0_ is the “natural” transmission rate, *α* a government-action strength parameter, *D* an additional category that measures the public’s reaction to the disease, and *N* the total population. However, the quality of the available data may not allow for a clean separation of government and public reactions. Furthermore, when the expression in Eq. (3) was applied to the Wuhan data, the fit parameter *κ* turned out to be very large, (*κ* > 1000), suggesting poor transferability from country to country. An additional difficulty with Eq. (3) is that the fit parameters will depend on the reporting ratio in a non-trivial way. Accordingly, the simpler expression

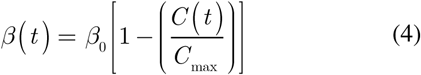

is used here. The parameter *C*_max_ corresponds to the maximum number of cases, and reflects the combined effort of government and society at large. A significant of using Eq. (4) is that *β*_0_ becomes independent of advantage underreporting if *C*_*max*_ is scaled according to the underreporting factor. Therefore, one can ignore underreporting when it comes to modeling the data.

The delay differential equation for *I*(*t*) in Eq (2) becomes an ordinary differential equation when solved between times *nτ* and (*n* + 1)*τ* using values of *I*(*t*) previously calculated between (*n* − 1)τ and *nτ*. For the first interval, values at t < 0 are needed. These are taken from the available data or approximated as *C*(*t*) = *I*(*t*) =*C*_0_ exp (*β* _0_ *t*)

It is apparent that Eq. (2) will not apply to times during which zoonotic transmission is ongoing and community spread is not generalized. During this period the evolution of the disease depends on random events and geographical peculiarities that cannot be modeled without the introduction of several more parameters and categories. To avoid these complications, the initial time *t* = 0 is taken as the time at which the total number of cases *C* has reached 500, and the data is fitted for *t* > 0 using *β*_0_, *C*_max_ and *τ* as adjustable parameters. Since at earlier times *C* ∼ *I*, one can further simplify Eq. (2) to *C*′(*t*) = *β*(*t*) *C*(*t*) and adjust the solution to the data using only *β*_0_ and *C*_max_ as adjustable parameters. In a subsequent stage of the fit, the solution can be extended to longer times by adjusting the time *τ*. The dashed lines in Figure 1 and 2 show the fits for China and South Korea, respectively, and the corresponding parameters are on Table I. For China, there is a well-known discontinuity in the reported data that makes the fit somewhat challenging. For South Korea, *C* ^*^ (*t*) seemed to be growing linearly as a function of time as this paper was written. This behavior is not captured by Eq. (4), and the fit “reacts” by yielding a higher value of *β*_0_ relative to China and an adjusted value of *C*_max_ that is somewhat smaller than the total number of cases already reported. This could be fixed by assuming that the transmission rate in Eq. (4) approaches an asymptotic residual value rather than becoming exactly zero. On the other hand, the total number of cases does seem to saturate to a maximum value in China, in nearly perfect agreement with Eq. (4). Accordingly, the origin of the late linear growth in South Korea may be an artifact, or a consequence of the discovery of new cases due the reportedly unrelenting COVID-19 testing in this country. Be it as it may, it seems premature at this time to refine the model to account for these effects.

**TABLE I.** Fit parameters of the time-delay COVID-19 model in Eq. (2) and (4). The last three columns correspond to the two-convalescence times extension in Eq. (5) The numbers in parenthesis indicate fit errors. For convalescence times, the error is about 1 day.

| Country | $\beta_0$<br>(day<sup>-1</sup>) | $C_{\max}$ | $\tau$<br>(day) | $\alpha$ | $\tau_1$<br>(day) | $\tau_2$<br>(day) |
| --- | --- | --- | --- | --- | --- | --- |
| China | 0.222(6) | 80800(400) | 25 | 0.4(1) | 17 | 34 |
| S. Korea | 0.30(1) | 8560(80) | 30 | 0.2(1) | 25 | 33 |
| Italy | 0.19(1) | 125000(5000) | 18 |  |  |  |
| Spain | 0.22(1) | 160000(30000) | 17 |  |  |  |

The time-delay model predicts correctly that the ratio *I*(*t*_peak_) / *C*(*t*_peak_) will be very close to unity. However, in the case of China the actual *I*(*t*) curve is only in qualitative agreement with the data. This implies that the approximation of a single convalescence time *τ* is too crude. In principle, one should use a distribution of convalescence times, but one can obtain a reasonable good agreement with the available data using two convalescence times *τ*_1_ and *τ*_2_, so that the second equation in (2) becomes

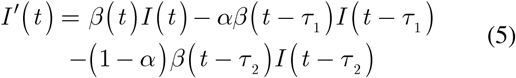

where 0 ≤ *α* ≤ 1.

The fit with the two convalescence times is shown as a solid line in Figure 1, and the corresponding parameters appear in Table I. It is apparent that the agreement is much improved. A two-convalescence time fit for South-Korea also leads to a small but noticeably improvement. The meaning of the new fit parameters, however, is not obvious. According to the Report of the WHO-China Joint Mission on Coronavirus Disease 2019 (COVID-19) [9], about 80% of the patients experience a mild disease and recover faster. However, it seems impossible to obtain a good fit using *α* = 0.80 if *τ*_1_ < *τ*_2_. This may indicate that the documented cases include a smaller fraction of mild cases, which would go mostly undetected. On the other hand, this finding may simply indicate that the two-step survival function implied by Eq. (5) still too naïve.

The model presented here, although extremely simplified, could be useful to health authorities by predicting the total number of cases as well as the peak number of infected people. The fit values could be improved on a daily basis as more data become available. As an example, Fig. 3 shows fits for Italy and Spain, which have not yet reached *t*_peak_ but show clear signs of a flattening in the number of cases curve. For other countries, the model does not yet provide a meaningful fit. In the case of the US, the growth as of March 24, 2020 is faster than exponential, perhaps as a result of improved testing. Iran, on the other hand, is a notable case because *C*\* (*t*) is quite linear, a behavior that the model here or any SEIR model fail to reproduce.

**FIG. 3.**
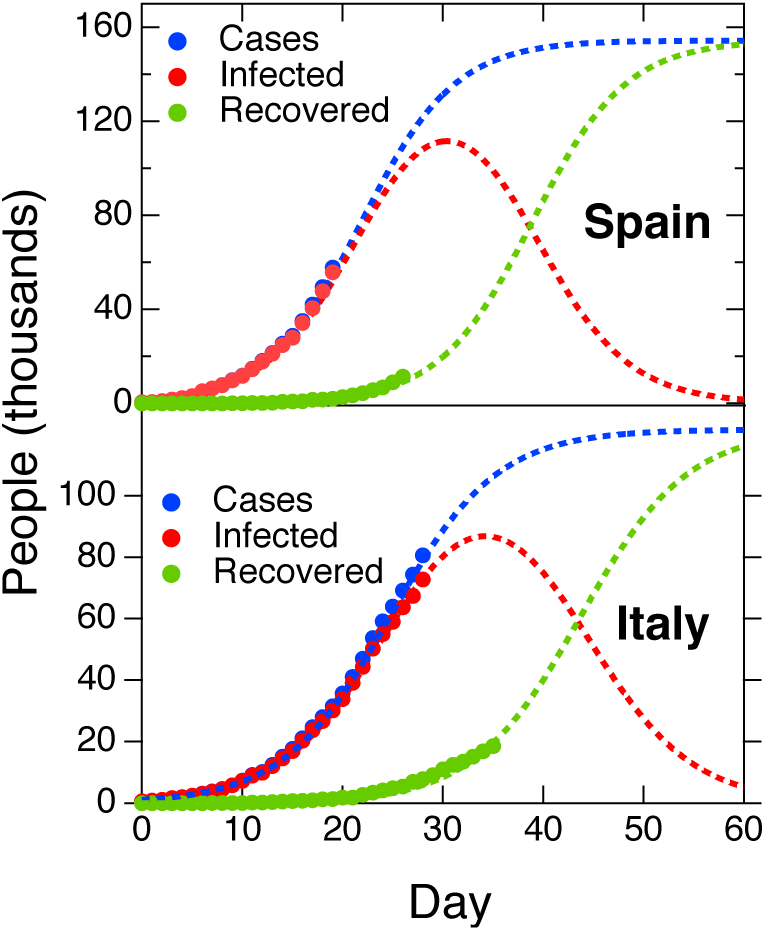
COVID-19 in Italy and Spain: blue dots show the number of cumulative cases, red dots the number of infected people at any given time, and green dots the cumulative number of recovered individuals. The data have been corrected according to Eq. (1) using *T* = 7 days. Day 0 is February 20, 2020 for Italy, and February 29, 2020 for Spain. The dotted lines of the same color represent the prediction from Eq. (2) with parameters from Table I. The solid line is the prediction using Eq. (5).

In summary, an elementary model of COVID-19 dynamics has been presented that produces a much better fit of existing data than standard SEIR models. The model is based on time-delay differential equations that take into account the convalescence time *τ*. It fits very well existing data for China and S. Korea, and it can be used to make predictions for several countries that have not yet reached the peak of the infected curve.

The numerical simulations and fits shown here were carrier out on IGOR PRO 8.0 (Wavemetrics, Inc). The code is available upon request.

## Data Availability

Analyzed data is publicly avaialable from numerous databases and even reputable newspapers. The code used to analyzed the data is described in the manuscript and freely available from the author upon request.

